# Trends and Differences in Breast Cancer Mortality by Race, Region and Age among Brazilian Women between 1996-2024: A Nationwide Ecological Study

**DOI:** 10.64898/2026.09.18.26363058

**Authors:** Angela Theresa Zuffo Yabrude, Leila Caroline Souza Reis, Ana Elisa Saragossa Zafalon, Clara Fish Araujo de Albuquerque, Eliza Del Fiol Manna

**Affiliations:** Universidade Regional de Blumenau (FURB), Blumenau, Brazil; University of São Paulo, São Paulo, Brazil; Barretos Cancer Hospital (Hospital de Amor), Barretos, São Paulo, Brazil; Università degli studi di Bari Aldo Moro, Bari, Italy; Unimed Sorocaba, Sorocaba, Brazil

**Keywords:** Breast cancer, Mortality, Health disparities, Time trends, Brazil

## Abstract

**Objectives:** To quantify the demographic contribution to Brazil’s rising number of breast cancer deaths and compare mortality trends by region and race/skin color.

**Study design:** Nationwide, population-based ecological time-series study.

**Methods:** Deaths from breast cancer (ICD-10 C50) in the Brazilian Mortality Information System, 1996-2024, were analysed. Crude rates use two denominators, the whole female population and women aged 20 and over. Age-standardized rates used the 2024 IBGE projection and, by race/skin color, Census populations for 2010 and 2022. Trends used weighted log-linear regression with BIC-selected joinpoints, and deaths were decomposed by the Das Gupta method.

**Results:** Deaths rose from 7,085 (1996) to 20,849 (2024). Over 2000-2024 the age-standardized rate rose from 11.4 to 14.3 per 100,000 women, with one joinpoint at 2017 (average annual percent change +0.87%, 95% CI 0.77 to 0.96). Growth and ageing accounted for 74.3% of the 12,542 additional deaths at ages 20 and over. Standardized rates rose in every region, fastest in the North (+3.09%/year) and slowest in the Southeast (+0.24%/year), narrowing the highest-to-lowest ratio from 2.67 to 1.38. White women had the highest age-standardized rate at both censuses (2022: 15.96 versus 12.28 and 11.48 per 100,000), Brown women had the larger increase (ratio of relative changes 1.20, 1.13 to 1.26).

**Conclusions:** Population growth and ageing accounted for most of the increase in the number of recorded breast cancer deaths at ages 20 and over. Regional differences in level narrowed, White women had the highest age-standardized rates at both censuses and Brown women the larger increase.

## 1. INTRODUCTION

Despite advances in diagnosis and treatment, breast cancer remains the leading cause of cancer death among Brazilian women, accounting for more than 20,000 deaths in 2023(1). These deaths occur within a universal public health system (SUS) serving a population of continental dimensions and marked social and regional heterogeneity, in which the absolute number of breast cancer deaths has risen steeply. Whether this rise reflects an increase in mortality risk or, in large part, the growth and ageing of the female population cannot be determined from absolute counts alone, a distinction of relevance given that breast cancer accounts for the largest cancer-related productivity loss among women in Brazil(2).

Breast cancer mortality in Brazil is neither socially nor geographically uniform. Social determinants, including socioeconomic position, educational attainment, geographic access to health services, and structural racial inequities, influence stage at diagnosis, treatment access, and survival outcomes across Brazil(3, 4). Mortality has historically been highest in the wealthier South and Southeast regions, a pattern associated with regional differences in socioeconomic conditions and health-service availability(5). Disparities by race/skin color recorded on the death certificate following the official IBGE classification are well documented: Black and Brown (mixed-race) women present at more advanced stages(6) and have poorer survival, as do women of lower socioeconomic status(7).

How these differences have evolved nationally remains incompletely characterized. Previous Brazilian studies have examined geographic and racial differences in breast cancer incidence, mortality and survival(6–9), mostly for single cities or states or for one dimension at a time. The present study integrates age-standardized regional trends, a formal decomposition of the change in the number of deaths and Census-based race-specific rates.

## 2. METHODS

### 2.1. Study design and data sources

This is a nationwide, population-based ecological time-series study. The numerator comprised all deaths in women with an underlying cause of malignant neoplasm of the breast (CID-BR-10 group 041; ICD-10 C50), by year of death from 1996 to 2024 and by place of residence, recorded in the Mortality Information System (SIM) and extracted through the public TabNet interface (DATASUS)(10) in February 2026, when the extraction reported the series as final through 2024. Deaths were extracted both as single-variable distributions and as joint cross-tabulations of region by age by year, race by age by year, and schooling by age. Of the 380,037 records, 57 were of women below 20 years and 88 had unknown age. Both were excluded rather than redistributed, leaving 379,892 deaths, 99.96% of the total. Denominators are the female population by five-year age group for Brazil and each region, 2000-2024 (IBGE Projection of the Population, 2024 revision), and by race/skin color and age from the 2010 and 2022 Censuses (IBGE, SIDRA table 9606).

### 2.2. Mortality rates and age standardization

We computed crude (CMR) and age-standardized (ASMR) mortality rates per 100,000 women. Crude rates are reported on two denominators, the whole female population and women aged 20 and over, and the denominator is stated wherever a crude rate appears, regional crude rates use the 20 and over denominator, so that they share the universe of the standardized regional series. ASMR was obtained by the direct method using the WHO World Standard Population (11), numerator age bands were harmonized to the denominator bands, and deaths with unknown age were excluded rather than redistributed. The standard weights were normalized over the full age range of the standard rather than within the 20 and over range, so the resulting rate is on the scale of an all-age standardized rate and is not a rate standardized within the population aged 20 and over, the conversion between the two scales is given in the Supplementary Methods(9). 95% confidence intervals for the ASMR were derived from the Fay-Feuer gamma method(12). Because population denominators were available from 2000, rate analyses were restricted to 2000-2024, count- and age-at-death analyses used the full 1996-2024 series, and race-stratified rates use the 2010 and 2022 Censuses.

### 2.3. Trend analysis

Temporal trends were summarized by the annual percent change (APC), estimated from inverse-variance-weighted log-linear regression of the natural logarithm of the rate on calendar year, with weights equal to the number of deaths, the average annual percent change (AAPC) was computed over the full period. For a standardized rate that weighting is an approximation, refitting with the exact inverse of Var[log(ASMR)] reproduced the estimate to three decimals, and the unweighted estimator and a Prais-Winsten AR(1) model are also reported. To assess changes in slope, a joinpoint model (0-2 joinpoints, continuous piecewise log-linear) was selected by the Bayesian Information Criterion, computed on the weighted residual sum of squares with 2K+2 parameters, K being the number of joinpoints(13), candidate locations were enumerated by exhaustive grid search, requiring at least five whole years between consecutive segment boundaries. A sensitivity analysis excluded the 2020-2021 pandemic years. The demographic contribution was quantified by a Das Gupta symmetric decomposition of deaths at 20 years and over into population size, age structure and age-specific rates (14).

### 2.4. Subgroup and age-at-death analyses

We estimated regional age-standardized rates for every year from 2000 to 2024, with crude rates on the same age domain alongside, and age-specific rates with their APCs, and the proportional distribution of deaths by region, race/skin color and schooling over time. Race-specific standardized rates were computed for 2010 and 2022 using Census female population by race and age. Deaths with unknown race/skin color were redistributed proportionally within each age band across the five recorded categories, observed and redistributed counts are both shown, and the redistributed totals reproduce the observed totals plus the eligible unknown records exactly. Age-adjusted rate ratios came from Poisson regression of death counts on race and age band with log population as offset and White women as reference, fitted to each census year with a race-by-age interaction, and the change between censuses was summarized by the ratio of relative changes. Where population denominators were unavailable (schooling), we analyzed the age at death using five-year band midpoints.

### 2.5. Software and reporting

Analyses were performed in R 4.5.3. Numerator-denominator reconciliation across the joint extractions was exact: the death totals obtained by race and by region agree in all 29 years. The study is reported in accordance with the STROBE statement (15).

### 2.6. Ethics

The study used publicly available, aggregate, de-identified secondary data, under Brazilian National Health Council norms governing research with public, anonymized information, institutional review board approval and informed consent were not required.

## 3. RESULTS

### 3.1. Overall mortality

Breast cancer deaths in Brazilian women increased from 7,085 in 1996 to 20,849 in 2024, a 2.9- fold rise. Over 2000-2024 the crude mortality rate climbed from 9.34 to 19.14 per 100,000 on the whole female population (APC +3.03%/year, 95% CI 2.91 to 3.14) and from 15.38 to 25.71 among women aged 20 and over (+2.19%/year, 2.08 to 2.29), whereas the age-standardized rate rose far more modestly, from 11.38 to 14.27 per 100,000 (APC +0.89%/year, 95% CI 0.78 to 1.00) (Fig. 1, Table 1, Supplementary Table S1). Of the 12,542 additional deaths among women aged 20 years and over between 2000 and 2024, 5,503 (43.9%) were attributed to population growth, 3,816 (30.4%) to ageing and 3,222 (25.7%) to changes in age-specific recorded mortality rates (Table 2; Figure 2). The BIC-selected model identified one joinpoint at 2017: the APC was +1.04%/year (95% CI 0.89 to 1.19) during 2000-2017 and +0.44%/year (0.10 to 0.79) during 2017-2024, yielding an AAPC of +0.87% (0.77 to 0.96) (Supplementary Table S2). The positive direction and overall magnitude of the national trend were consistent across sensitivity analyses (Supplementary Table S3, Supplementary Figure S1).

**Figure 1.**
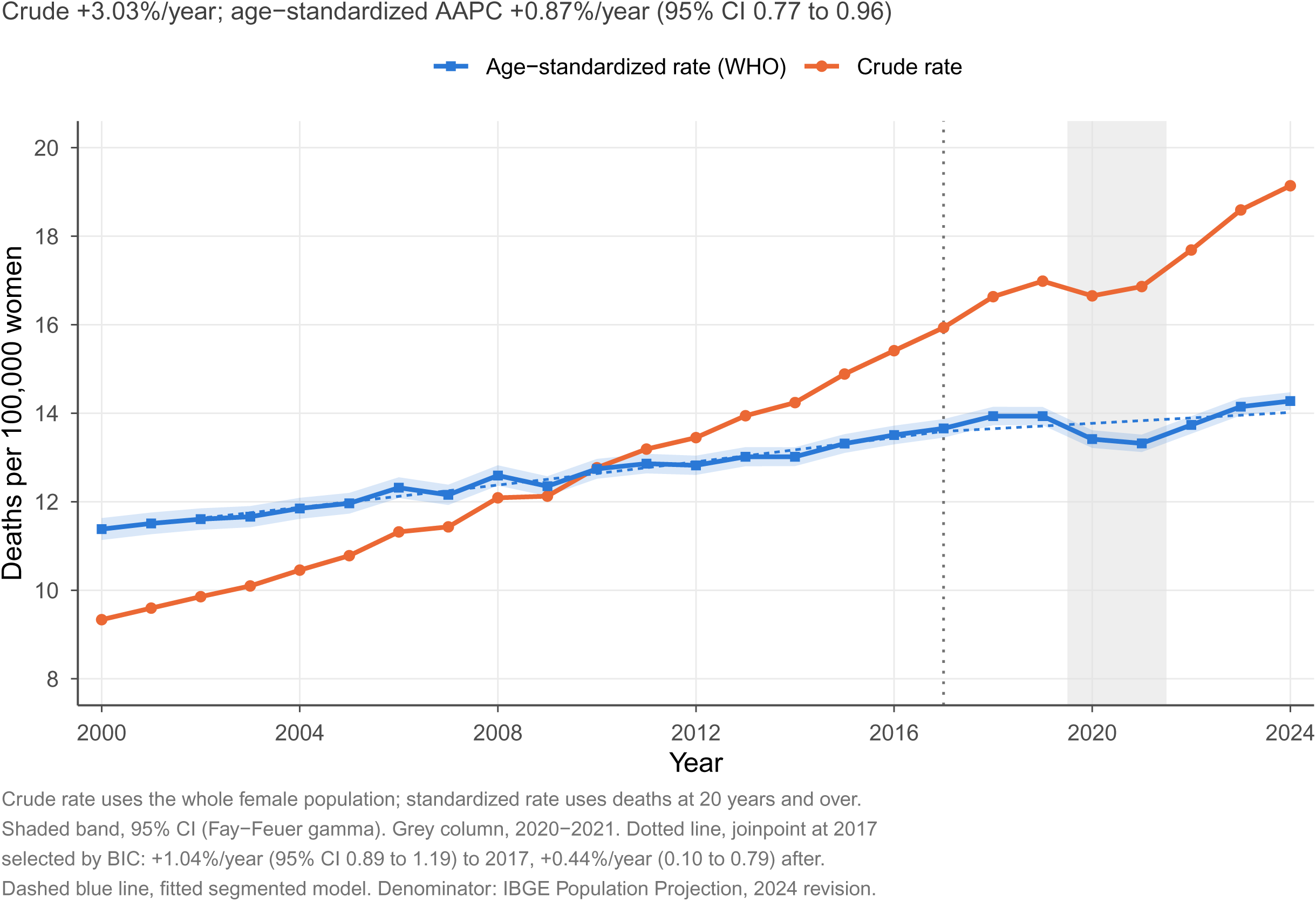
Crude and age-standardized breast cancer mortality in Brazilian women, 2000-2024. The crude rate uses the whole female population, and the standardized rate uses deaths at 20 years and over; shaded band, 95% CI (Fay-Feuer). Dotted line, joinpoint at 2017; dashed line, fitted segmented model; shaded column, 2020-2021.

**Figure 2.**
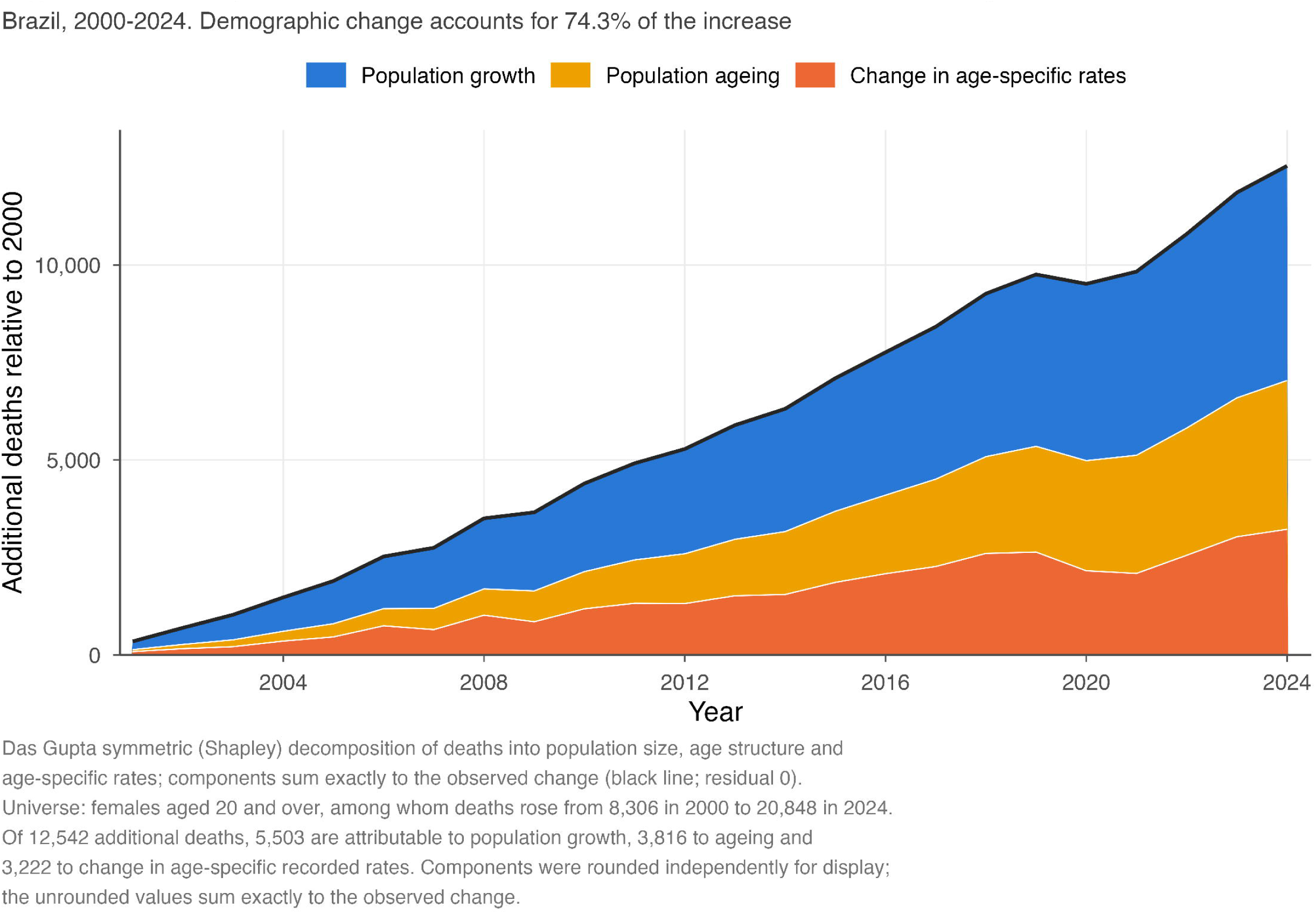
Das Gupta decomposition of the increase of 12,542 breast cancer deaths among women aged 20 years and over, Brazil, 2000-2024, into population growth, population ageing and change in age-specific recorded rates. Components were rounded independently for display; the unrounded values sum exactly to the observed change of 12,542 deaths.

**Table 1.** Crude and age-standardized breast cancer mortality rates and annual percent change, Brazil and regions, 2000-2024, with the segmented analysis of the national age-standardized series. Rates per 100,000 women. The age domain of each row is stated in the row label: the national crude rate is given on the whole female population and on women aged 20 and over, and regional crude rates use the 20 and over denominator so that they share the universe of the standardized series. Age-standardized rates use deaths at 20 years and over with WHO World Standard weights normalized over the full age range of the standard (see Methods). Annual percent change from log-linear regression weighted by the number of deaths. Denominators: IBGE Projection of the Population, 2024 revision. AAPC, average annual percent change.

| Stratum | Rate 2000 | Rate 2024 | APC, %/yr | 95% CI |
| --- | --- | --- | --- | --- |
| Brazil, crude rate, whole female population | 9.34 | 19.14 | 3.03 | 2.91 to 3.14 |
| Brazil, crude rate, women aged 20 and over | 15.38 | 25.71 | 2.19 | 2.08 to 2.29 |
| Brazil, age-standardized rate (WHO) | 11.38 | 14.27 | 0.89 | 0.78 to 1.00 |
| Region, age-standardized: North | 5.35 | 11.36 | 3.09 | 2.71 to 3.47 |
| Region, age-standardized: Northeast | 6.48 | 13.16 | 2.51 | 2.10 to 2.91 |
| Region, age-standardized: Central-West | 9.80 | 13.46 | 1.43 | 1.07 to 1.78 |
| Region, age-standardized: South | 14.25 | 15.65 | 0.43 | 0.30 to 0.57 |
| Region, age-standardized: Southeast | 14.02 | 14.95 | 0.24 | 0.14 to 0.35 |
| Region, crude (20+): Central-West | 11.51 | 22.23 | 3.02 | 2.69 to 3.36 |
| Region, crude (20+): Northeast | 8.76 | 22.72 | 3.68 | 3.34 to 4.02 |
| Region, crude (20+): North | 6.07 | 16.50 | 4.34 | 3.98 to 4.69 |
| Region, crude (20+): Southeast | 19.56 | 28.42 | 1.61 | 1.49 to 1.73 |
| Region, crude (20+): South | 19.77 | 29.84 | 1.80 | 1.66 to 1.94 |
| Joinpoint analysis of the national age-standardized series |  |  |  |  |
| Segment 2000-2017 |  |  | 1.04 | 0.89 to 1.19 |
| Segment 2017-2024 |  |  | 0.44 | 0.10 to 0.79 |
| AAPC 2000-2024 |  |  | 0.87 | 0.77 to 0.96 |

**Table 2.** Das Gupta decomposition of the change in the number of breast cancer deaths among Brazilian women aged 20 years and over, 2000-2024. Components are symmetric (Shapley) averages over all orders of substitution. Unrounded components sum exactly to the observed change of 12,542 deaths, with a residual of 0.00e+00; the displayed values were rounded independently and may therefore not sum exactly to the total. Had the 2000 age-specific rates persisted, 16,504 deaths would have been expected in 2024; that counterfactual applies the 2000 rates to the 2024 age structure and is not identical to the rate component above.

| Component | Additional deaths | % of increase | Unrounded |
| --- | --- | --- | --- |
| Population growth | 5,503 | 43.9 | 5503.228 |
| Population ageing | 3,816 | 30.4 | 3816.439 |
| Change in age-specific rates | 3,222 | 25.7 | 3222.333 |
| Total observed change | 12,542 | 100.0 | 12542.000 |

### 3.2. Age

Age-specific mortality rates rose in every band and the trends differed across bands (quasi-Poisson age-by-year interaction, F = 56.9 on 6 and 161 degrees of freedom, p < 0.001): increases were steepest in women aged 80 years and over (APC +2.02%/year, 95% CI 1.86-2.19) and, proportionally, in younger women (20-29: +2.99%/year on 73 deaths in 2000 and 157 in 2024, 30-39: +1.88%/year), with the slowest change at 50-59 years (+0.36%/year) (Supplementary Figures S2 and S3; Supplementary Table S4). Because these are age-specific rates, the increase at 80 years and over cannot be explained solely by growth in the number of women in that age group, although the band is open-ended.

### 3.3. Race / skin color

Among deaths with valid race coding, the share occurring in Black and Brown women rose from 24.5% to 42.7% between 2000 and 2024, while records with missing race/skin color fell from 10.7% to 1.2%, under extreme bounds the increase cannot be explained by improved completeness alone (Supplementary Table S5, Supplementary Figure S4). Using Census female population by race and age, age-standardized mortality was highest in White women at both censuses: 15.38 per 100,000 (95% CI 15.05 to 15.71) in 2010 and 15.96 (15.66 to 16.27) in 2022, against 12.63 and 12.28 in Black and 9.25 and 11.48 in Brown women (Table 3, Figure 3). Age-adjusted rate ratios in 2022 were 0.77 (0.72 to 0.80) for Black and 0.72 (0.69 to 0.74) for Brown women, with a significant race-by-age interaction (p<0.001). Between censuses the rate increased more among Brown than White women (+24.1% versus +3.8%; ratio of relative changes 1.20, 1.13 to 1.26), whereas the change among Black women did not differ (−2.8%, 0.94, 0.86 to 1.02). These are two cross-sections, not a trajectory. Among White, Black and Brown women, excluding rather than redistributing records with unknown race/skin color reduced the age-standardized rates by approximately 5.7% in 2010 and 1.7% in 2022, without materially changing the comparative findings (Supplementary Table S6). From 30 years upward every age-specific rate ratio was below 1, at 20-29 years the estimates were imprecise (Supplementary Table S7; Supplementary Figure S5).

**Figure 3.**
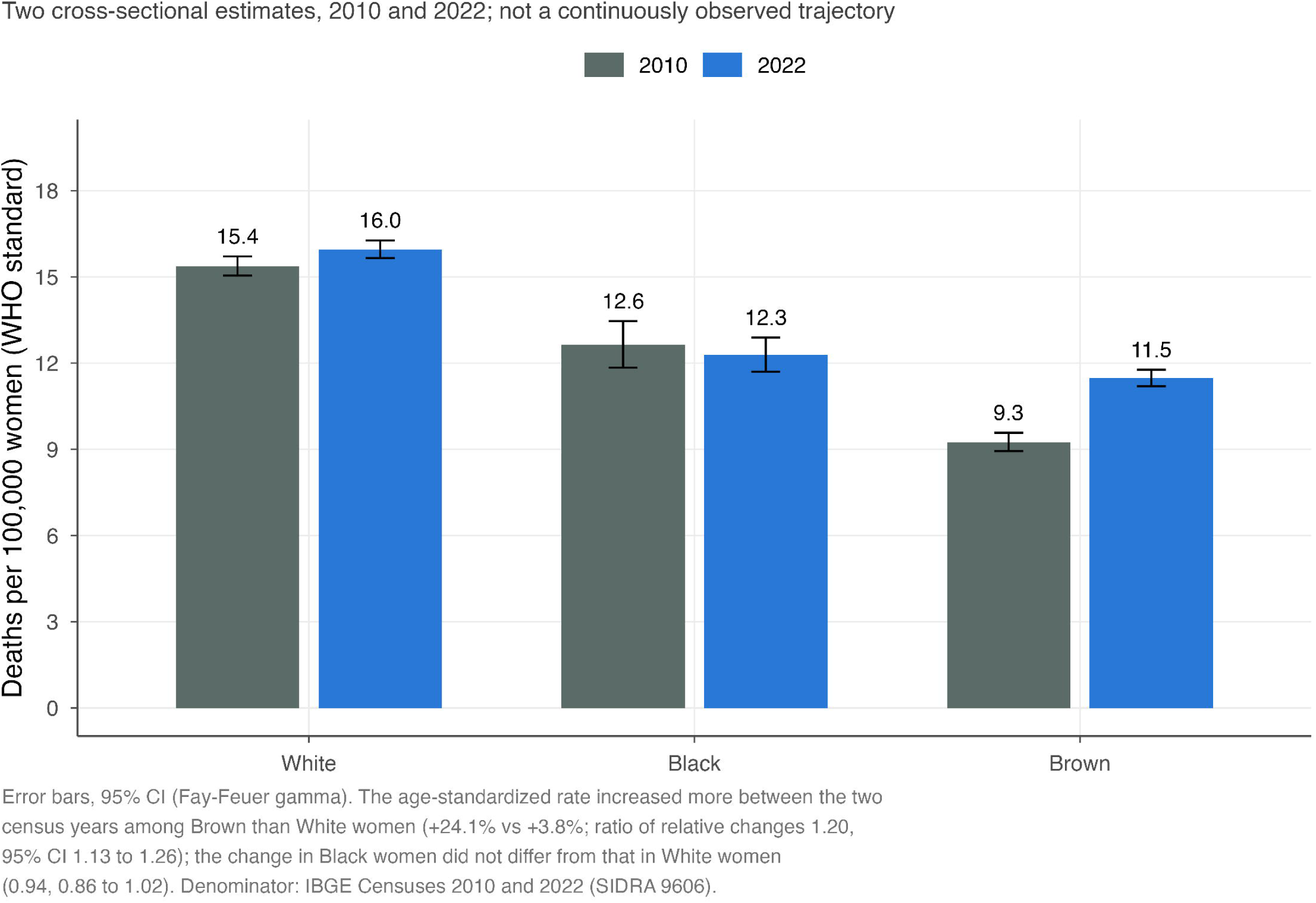
Age-standardized breast cancer mortality by race/skin color at the 2010 and 2022 Censuses, Brazil. Error bars, 95% CI (Fay-Feuer).

**Table 3.** Age-standardized breast cancer mortality by race/skin color, Brazilian Censuses of 2010 and 2022, and the change between them. Crude rates are per 100,000 women aged 20 years and over. Age-standardized rates use deaths at ages 20 and over with WHO standard weights normalized over the full age range; they are not rates standardized within the population aged 20 and over (see Methods). Observed deaths are the records as coded; the crude and standardized rates are computed after proportional redistribution of records with unknown race/skin color within each age band across all five recorded categories and therefore cannot be reconstructed from the observed counts alone. The redistributed totals reproduce the observed totals plus the eligible unknown records exactly (320 in 2022, 724 in 2010); Asian and Indigenous women take the remainder of the redistribution and are shown in Supplementary Table S6 rather than here, because of small numbers. 95% CI by the Fay-Feuer gamma method. Denominators: female population by race/skin color and age, Demographic Censuses 2010 and 2022 (IBGE, SIDRA table 9606*). Rate ratios are relative to White women. The ratio of relative changes is (ASMR2022/ASMR2010) in each group divided by the same ratio in White women; its interval comes from a Poisson contrast of the log relative changes and does not incorporate the uncertainty introduced by redistribution*.

| Year | Race/<br>Skin color | Observed<br>deaths | Redistribute<br>d deaths | Female<br>population | Crude<br>rate | ASMR | 95% CI | Rate<br>ratio |
| --- | --- | --- | --- | --- | --- | --- | --- | --- |
| 2010 | White | 7,875 | 8,347 | 33,549,213 | 24.88 | 15.38 | 15.05 to<br>15.71 | 1.000 |
| 2010 | Black | 900 | 955 | 5,125,451 | 18.63 | 12.63 | 11.84 to<br>13.46 | 0.821 |
| 2010 | Brown | 3,130 | 3,322 | 26,629,378 | 12.48 | 9.25 | 8.94 to<br>9.58 | 0.602 |
| 2022 | White | 10,934 | 11,118 | 35,323,750 | 31.48 | 15.96 | 15.66 to<br>16.27 | 1.000 |
| 2022 | Black | 1,644 | 1,672 | 8,052,197 | 20.77 | 12.28 | 11.70 to<br>12.89 | 0.770 |
| 2022 | Brown | 6,089 | 6,194 | 33,707,922 | 18.38 | 11.48 | 11.20 to<br>11.77 | 0.719 |

| Race/skin color | ASMR 2010 | ASMR 2022 | Relative<br>change | Ratio of<br>relative<br>changes | 95% CI |
| --- | --- | --- | --- | --- | --- |
| White | 15.38 | 15.96 | +3.8% | 1 (reference) |  |
| Black | 12.63 | 12.28 | -2.8% | 0.94 | 0.86 to 1.02 |
| Brown | 9.25 | 11.48 | +24.1% | 1.20 | 1.13 to 1.26 |

### 3.4. Region

Age-standardized regional rates converged from below: the North (+3.09%/year, 95% CI 2.71 to 3.47), Northeast (+2.51%, 2.10 to 2.91) and Central-West (+1.43%, 1.07 to 1.78) rose several times as fast as the South (+0.43%, 0.30 to 0.57) and Southeast (+0.24%, 0.14 to 0.35), climbing from low baselines toward the historically higher South/Southeast, so that the ratio between the highest and lowest regional rate fell from 2.67 in 2000 to 1.38 in 2024 (Figure 4; Table 1). Crude rates on the same age domain rose faster than the standardized rates in every region, from +4.34%/year in the North to +1.61% in the Southeast (Supplementary Figure S6), the difference being regional differences in age composition.

**Figure 4.**
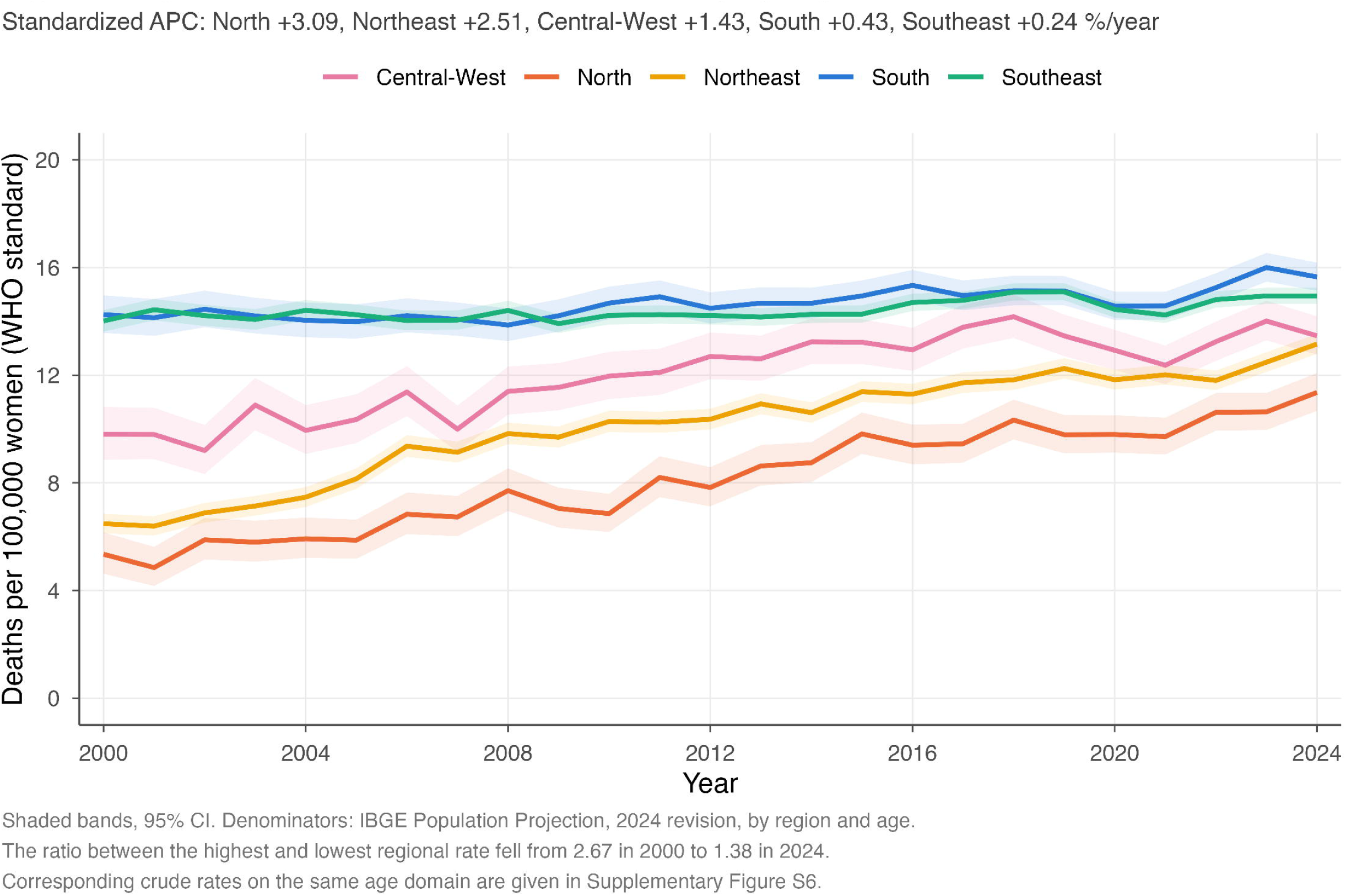
Age-standardized breast cancer mortality by region, Brazil, 2000-2024. Shaded bands, 95% CI. Denominators: IBGE Projection of the Population, 2024 revision, by region and age.

### 3.5. Schooling and age at death

Schooling and age at death describe the composition of recorded deaths rather than mortality risk. Among deaths of White, Black and Brown women aged 20 years and over in 2015-2024, mean age at death was 64.1, 61.0 and 59.5 years, respectively (Supplementary Table S8). Educational composition by age at death is presented descriptively in Supplementary Figure S7.

## 4. DISCUSSION

In this nationwide ecological time-series study, recorded breast cancer deaths rose almost threefold over nearly three decades, while the age-standardized rate rose far more modestly. The Das Gupta decomposition quantified that divergence: among women aged 20 and over, population growth and ageing accounted for 74.3% of the 12,542 additional deaths between 2000 and 2024. The remaining 25.7% is a change in recorded rates and not a direct measure of change in underlying risk, since recorded mortality also reflects diagnostic practice and death certification(17, 18).

The regional findings changed once denominators by region and age were used. Age-standardized mortality increased in all regions, with larger increases where baseline rates were lower, so the ratio between the highest and lowest regional rate narrowed from 2.67 to 1.38. The South and Southeast still had the highest standardized rates in 2024, consistent with greater diagnostic capacity and historically better death registration, while the North and Northeast include areas of greater socioeconomic vulnerability and lower service availability. Regional and racial composition overlap(19), so historically lower recorded rates there should not be read simply as lower risk(8, 17, 18).

Improvements in the Brazilian Mortality Information System, though essential for surveillance, can themselves raise recorded mortality where deaths were previously unregistered or assigned to ill-defined causes, and this matters most in the North and Northeast(17, 18). We applied no correction factors and cannot separate that contribution from change in underlying mortality. An independent analysis of the same source that did apply them raised the national number of deaths by 15.9% over 2000-2023, and by 67.2% in the North and 54.8% in the Northeast during 2000- 2007, falling steadily over time, because the under-registration repaired was concentrated in the early years, correction attenuated the estimated increase, to +0.28% nationally(9). Improvements in registration may therefore have inflated the increases we observe, particularly in the North and Northeast, the magnitude of that contribution was not estimated here.

The race and skin color results require the clearest restatement. With Census denominators, age-standardized mortality was highest in White women at both censuses, so the higher burden inferred from younger age at death does not hold as a statement about mortality rates. It is also consistent with higher incidence and with better diagnostic access and death registration where White women are concentrated. Two things differ at once: the level, higher in White women in both years, and the change between them, larger in Brown women. Elsewhere, mortality can exceed that of the majority population despite lower incidence(20). We cannot identify the mechanism here, since our data hold no information on stage, subtype, incidence or treatment(21–23).

Age at death and schooling describe the composition of recorded deaths rather than mortality risk. Black and Brown women died at younger ages than White women, the age-specific rates show this is not driven by higher rates at most ages, though they do not by themselves account for the whole distribution, which also reflects the size and age structure of each population at risk. The educational composition of deaths is compatible with generational differences in access to schooling.

The age-specific results carry two planning-relevant messages. The increase at 80 years and over cannot be explained solely by growth in the number of women in that age group, since the measure is an age-specific rate, although changes in the internal age composition of this open-ended band may contribute, that group will represent a growing share of the absolute burden. The increase at 20-29 years rests on very small numbers and matters mainly because deaths before age 50 carry high social and economic costs and often fall outside screening ages(24, 25). These findings place Brazil against the prevailing international trend: global age-standardized mortality has been falling even as incidence rises(26) and the United States death rate fell 44% between 1989 and 2022(20), while within Latin America Brazil sits in the rising group(27, 28). Absolute levels should be compared cautiously, because standard populations, age domains and mortality definitions differ.

The decline observed during 2020-2021 should be interpreted in the context of the COVID-19 pandemic. It is unlikely to represent a true improvement in breast cancer control. During the pandemic, screening, diagnostic procedures, elective care, and oncology services were disrupted, with documented differences by race/skin color(29). Mortality records may also have been affected by changes in health service use and cause-of-death certification. Excluding 2020 and 2021 from the segmented analysis moved the joinpoint by one year and left the average annual percent change essentially unchanged.

This study has several strengths. It used nationwide data spanning nearly three decades. National and regional standardized rates come from a single population source in which the national denominator equals the sum of the regional ones, the demographic contribution is quantified by a decomposition whose components sum to the observed change, and race-specific rates use Census denominators rather than an approximation.

The study also has limitations. It relied on secondary mortality data, which may be affected by underreporting, regional differences in data quality and changes in death certification over time, and we applied no correction factors(9, 17, 18). Denominators jointly stratified by race and age were available only for the two census years, so that comparison rests on two cross-sections and its intervals do not incorporate the uncertainty introduced by redistributing records with unknown race, no denominators exist for schooling or age at death, and both had missing or changing classifications. As an ecological study, it cannot infer individual risk. Finally, it lacked data on breast cancer incidence, stage at diagnosis, tumor subtype, screening history, treatment, and time to treatment.

The rising number of deaths reflects the demographic reality of an ageing population, while the age-standardized rate rises slowly and regional differences in level narrow. Brazil will need to prepare for a larger absolute burden and identifying why the differences we describe arise will require data on stage, treatment and access, including time to treatment(30), that mortality records do not contain.

## 5. CONCLUSIONS

Recorded breast cancer deaths in Brazil nearly tripled between 1996 and 2024. Over 2000-2024 the age-standardized rate rose from 11.4 to 14.3 per 100,000 women (AAPC +0.87%, 95% CI 0.77 to 0.96), and population growth and ageing accounted for 74.3% of the increase in the number of deaths at ages 20 and over. Standardized regional rates rose fastest in the North and slowest in the Southeast, narrowing the highest-to-lowest gap from 2.67-fold to 1.38-fold. At the 2010 and 2022 Censuses age-standardized mortality was highest in White women, while the increase between them was larger among Brown women. These are trends in recorded mortality, so the contrasts described are observed differences rather than established inequalities in underlying risk.

## Data Availability

All data produced in the present study are available upon reasonable request to the authors

## SUPPLEMENTARY MATERIAL

The following items are supplied as a separate Supplementary file, together with the aggregated data and the analysis scripts.

**Supplementary Table S1.** National crude rates on both denominators and age-standardized breast cancer mortality with 95% CI, Brazil, 2000-2024.

**Supplementary Table S2.** Joinpoint model selection: best model with zero, one and two joinpoints, with the weighted residual sum of squares and the Bayesian Information Criterion under both parameter conventions.

**Supplementary Table S3.** Sensitivity of the national trend estimate to the standard population, the weighting scheme, the error structure and exclusion of 2020-2021.

**Supplementary Table S4.** Age-specific breast cancer mortality rates and annual percent change by age band, Brazil, 2000-2024, with the unweighted estimator for comparison.

**Supplementary Table S5.** Completeness of race/skin color recording and extreme bounds for the Black and Brown share of breast cancer deaths, Brazil, 1996-2024.

**Supplementary Table S6.** Redistribution versus exclusion of deaths with unknown race/skin color at both censuses, for all groups with a Census denominator, with the resulting rate ratios and ratio of relative changes.

**Supplementary Table S7.** Age-specific rate ratios for breast cancer mortality by race/skin color, 2022 Census.

**Supplementary Table S8.** Age at death among women classified as White, Black or Brown (2015-2024) and by schooling (1996-2024) among recorded breast cancer deaths. Descriptive only; no population denominators are available for these variables.

**Supplementary Figure S1.** Sensitivity of the national annual percent change to the standard population, the weighting scheme, an AR(1) error structure and exclusion of 2020-2021. Error bars, 95% CI.

**Supplementary Figure S2.** Age-specific breast cancer mortality rates by ten-year age band, Brazil, 2000-2024, on a logarithmic scale.

**Supplementary Figure S3.** Annual percent change in age-specific breast cancer mortality rates by age group, Brazil, 2000-2024. Error bars, 95% CI.

**Supplementary Figure S4.** Completeness of race/skin color recording in breast cancer death certificates and extreme bounds for the Black and Brown share of deaths, Brazil, 1996-2024.

**Supplementary Figure S5.** Age-specific rate ratios for breast cancer mortality, Black and Brown versus White women, 2022 Census. Error bars, 95% CI.

**Supplementary Figure S6.** Crude breast cancer mortality by region, Brazil, 2000-2024, on the same age domain as the standardized series (women aged 20 and over).

**Supplementary Figure S7.** Educational composition of breast cancer deaths by age at death, Brazil, 2015-2024, among records with valid schooling.

## Tables and Supplementary Material

Trends and differences in breast cancer mortality by race, region and age among Brazilian women, 1996-2024. Every value below is read from a single analytical output file produced by the scripts supplied with this submission; none is entered by hand.

